# Factors influencing nursing students’ intention to accept COVID-19 vaccination – A pooled analysis of seven countries

**DOI:** 10.1101/2021.01.22.21250321

**Authors:** Evridiki Patelarou, Petros Galanis, Enkeleint A. Mechili, Agathi Argyriadi, Alexandros Argyriadis, Evanthia Asimakopoulou, Stiliana Brokaj, Jorgjia Bucaj, Juan Manuel Carmona-Torres, Ana Isabel Cobo-Cuenca, Jakub Doležel, Stefano Finotto, Darja Jarošová, Athina Kalokairinou, Daniela Mecugni, Velide Pulomenaj, Aurela Saliaj, Idriz Sopjani, Majlinda Zahaj, Athina Patelarou

## Abstract

Experiencing the second wave of COVID-19 pandemic, high vaccination coverage by a safe and effective vaccine globally would be a great achievement. Acceptance of vaccination by healthcare students is an important issue as they have a key role as future professionals in educating patients, informing and guiding them to the right clinical decision. The aim of this study was to explore the intention of nursing students to get vaccinated for SARS-CoV-2 infection and the factors acting either as motivators or barriers towards vaccination. A multicenter cross-sectional study was conducted in 7 countries (Greece, Albania, Cyprus, Spain, Italy, Czech Republic and Kosovo) through a web survey. In total 2249 undergraduate nursing students participated. Forty three point eight percent of students agreed to accept a safe and effective COVID-19 vaccine, while the acceptance was higher among Italian students. The factors for intention to get vaccinated were male gender (p=0.008), no working experience in healthcare facilities during the pandemic (p=0.001), vaccination for influenza in 2019 and 2020 (p<0.001), trust in doctors (p<0.001), governments and experts (p=0.012), high level of knowledge (p<0.001) and fear of COVID-19 (p<0.001). Understanding of factors that influence students’ decision to accept COVID-19 vaccination could increase the acceptance rate contributing to a management of the pandemic.

**Highlights:**

- Less than half of the sample intended to accept COVID-19 vaccination
- Factors that influenced nursing students to get vaccinated against COVID-19 were male gender, no working experience in healthcare facilities during the pandemic, vaccination for influenza in 2019 and 2020, trust in doctors, governments and experts, high level of knowledge and fear of COVID-19.

## Introduction

SARS-CoV-2 (COVID-19) infection was declared as a pandemic by the World Health Organization (WHO) on March 12^th^ 2020 [1]. The second wave of COVID-19 pandemic is now hitting European countries with the total number of confirmed cases exceeding 87 million and 1.9 million deaths until now (January 2021) [2]. The impact of the pandemic is far more than a health crisis as it is affecting society and economy and although it varies from country to country it will definitely increase poverty, unemployment, social distancing, self-isolation, inequalities globally with serious psychosocial impact [3,4,5]. As a result, responding swiftly to the pandemic and beat COVID-19 has now become the most important thing for humanity. It is believed that with the availability of a safe and effective vaccine for COVID-19, high vaccination coverage globally will be achieved, and a great progress will be made in controlling the pandemic [6].

Towards this direction, studies on COVID-19 vaccine are ongoing and several vaccines have already been launched into the market for the control of COVID-19. However, vaccines availability doesn’t guarantee population vaccination due to the increase of antivaccination movement and vaccine hesitancy which consists the next challenge in the fight against COVID-19 [7]. Vaccine hesitancy is a chronic public health threat that may undermine efforts to achieve herd immunity by vaccination [8]. Despite the overwhelming volume of evidence on the benefits of immunization, widespread misconceptions and mistrust of information about vaccine efficacy and safety remain [8]. Key barriers to vaccination include lack of knowledge and confidence, lack of access to vaccines, concerns about the efficacy and safety of vaccines, and religious beliefs [9]. These barriers are also empowered by different conspiracy theories that circulate mainly in the social media.

Surprisingly, vaccine hesitancy phenomenon is present even among healthcare professionals. Vaccination of healthcare professionals is of utmost importance to prevent the spread of viruses as they are in the best position to understand patients doubts and concerns, to respond to their questions, and to explain in simple words to them the importance and positives of vaccination [10]. However, more and more studies report low acceptance levels and high hesitancy level to COVID vaccination among healthcare professionals (medical doctors, nurses, dentists etc.), including those who provide vaccination to patients [9, 11]. Almost all of these studies focused on healthcare professionals’ attitudes and concerns related to insufficient knowledge, efficacy and effectiveness of the vaccine, and its potential long-term side effects. In addition, acceptance of vaccination by healthcare students is of paramount importance as they act as information providers to patients and they will also be the future professionals. Existing studies on vaccine hesitancy among healthcare students are rare and no previous studies have examined the extent at which vaccine hesitancy phenomenon is evident in this specific population. We therefore aimed to explore the intention of nursing students to get vaccinated for SARS-CoV-2 infection and the key factors acting either as motivators or barriers towards vaccination.

## Methodology

### Design and participants

A multicenter cross-sectional study was conducted in seven countries (Greece, Albania, Cyprus, Spain, Italy, Czech Republic and Kosovo) during the so-called second wave of the COVID-19 pandemic. Particularly, in all countries data was collected during December 2020. Our sample consisted of undergraduate nursing students who were attending classes through online or face-to-face classes organized by 7 universities in participating countries (Greece-Hellenic Mediterranean University, Albania-University of Vlora, Cyprus-Frederick University, Spain-University of Castilla-La Mancha & University of Toledo, Italy-University of Modena and Reggio Emilia, Czech Republic-University of Ostrava, Kosovo-AAB College). We included in total 2249 participants from Kosovo (n=1020), Albania (n=313), Greece (n=275), Spain (n=181), Italy (n=170), Czech Republic (n=159), and Cyprus (n=131).

### Procedure

Students were asked to participate in the study through a web survey which included general information regarding the purpose and the process of the study and the informed consent as well. Ethical committees of all participated universities have reviewed and approved the study protocol. No personal data was recorded, the questionnaire was anonymous and an informed consent was obtained at the beginning of the questionnaire from each student confirming his or her willingness to participate.

The questionnaire comprised 29 items, which take about 6-8 minutes to complete. The first part of the study questionnaire included questions about demographic characteristics, perceived knowledge and beliefs regarding coronavirus and COVID-19 vaccine, trust towards the experts, doctors and government and factors influencing the students’ intention to vaccinate against the COVID-19 virus. The questions of the first part were developed initially in English, translated, and adapted in local languages by the local research team. The second part of the instrument included the Fear of COVID-19 Scale (FCV-19S) which was used to measure the fear against the coronavirus [12]. FCV-19S is a self-reported scale which comprises seven items rated on a 5-point Likert-type scale (1 = strongly disagree to 5 = strongly agree). A total individual score can be calculated ranging from 7 to 35 with higher scores representing higher level of fear against the coronavirus disease. FCV-19S has been translated and validated into Greek, Spanish and Italian and has been used in previous studies to measure fear of COVID-19 levels [13-15]. For the purposes of the present study, the Albanian and Czech research teams translated and validated the FCV-19S in their local language. The internal consistency for FCV-19S was excellent (Cronbach’s alpha=0.87).

### Statistical analysis

Continuous variables are presented as mean, standard deviation, median, and range, while categorical variables are presented as numbers (percentages).

We considered demographic data and students’ answers regarding the COVID-19 and vaccination as the independent variables and intention to accept COVID-19 a safe and effective vaccine as the dependent variable.

We converted age in a dichotomous variable according to median value. Responses that were recorded on a five-point Likert scale (“completely disagree”, “somewhat disagree”, “neutral”, “somewhat agree” and “completely agree”) were categorized in three categories; disagree (“completely disagree” and “somewhat disagree”), neutral, and agree (“somewhat agree” and “completely agree”). We used quantiles to convert the score on fear of COVID-19 scale into an ordinal variable with five categories; very low fear, low, medium, high, and very high fear. Since there were data from seven countries, we grouped them according to the deaths per million population. For deaths per million population (mortality), we categorized the continuous values into categories of low (fewer than 400 deaths per million population), medium (between 400 and 800 deaths per million population) and high (more than 800 deaths per million population [16]. Low mortality group included Cyprus, medium mortality group included Greece, Kosovo and Albania, and high mortality group included Spain, Czech Republic and Italy.

We used multivariable logistic regression to eliminate confounding. In that case, we defined the outcome as 1 if a participant answered “somewhat agree” or “completely agree” and 0 for any other response. First, we conducted univariate logistic regression analysis and then variables that were significantly different (p<0.20) were entered into the backward stepwise multivariate logistic regression analysis. We estimated adjusted odds ratios (OR) with 95% confidence intervals and p-values.

All tests of statistical significance were two-tailed, and p-values<0.05 were considered significant. Statistical analysis was performed with the Statistical Package for Social Sciences software (IBM Corp. Released 2012. IBM SPSS Statistics for Windows, Version 21.0. Armonk, NY: IBM Corp.).

### Ethical issues

All ethical issues were followed during the study. Participation was voluntary and participants could withdraw at any moment. No personal data was recorded. Participants were assured that all data collected will be used only for the current study. All Universities ethical committees approved the study. Additionally, before completing the questionnaire, participants were asked to give their consent to participate in the study. Approval was also received from the developer of FCV-19S instrument.

## Results

Study population included 2249 nursing students in total and demographic characteristics of the students according to the country residence are shown in Table 1. The 84.7% (n=1902) of the students were of female gender and 88.6% declared single. During the COVID-19 pandemic, more than a third of the students (40.3%) with at least one person belonging at vulnerable groups (elderly, chronic condition), 33.6% had clinical practice in healthcare facilities, and 14.2% worked in healthcare facilities.

**Table 1.** Demographic characteristics of the students.

| Characteristics | N | % |
| --- | --- | --- |
| Gender |  |  |
| Male | 344 | 15.3 |
| Female | 1902 | 84.7 |
| Age (years), mean, standard deviation | 21.6 | 5.6 |
| Country of origin |  |  |
| Greece | 275 | 12.2 |
| Spain | 181 | 8.0 |
| Cyprus | 131 | 5.8 |
| Czech Republic | 159 | 7.1 |
| Italy | 170 | 7.6 |
| Albania | 313 | 13.9 |
| Kosovo | 1020 | 45.4 |
| Marital status |  |  |
| Single | 1978 | 88.6 |
| Married | 233 | 10.4 |
| Widowed | 4 | 0.2 |
| Divorced | 18 | 0.8 |
| Chronic disease |  |  |
| Yes | 136 | 6.1 |
| No | 2110 | 93.9 |
| Living with vulnerable groups during the COVID-19 pandemic |  |  |
| Yes | 906 | 40.3 |
| No | 1341 | 59.7 |
| Clinical practice in healthcare facilities during the COVID-19 pandemic |  |  |
| Yes | 752 | 33.6 |
| No | 1486 | 66.4 |
| Working in healthcare facilities during the COVID-19 pandemic |  |  |
| Yes | 320 | 14.2 |
| No | 1926 | 85.8 |

Students’ answers regarding the COVID-19 and vaccination are listed in Table 2. Regarding the COVID-19 disease, 50.9% has been contacted with a confirmed or a suspected case, 11.8% has been infected and 60.4% has at least a family member/friend that has been infected. The 60% reported to have high to very high self-perceived knowledge about the COVID-19 and 54.9% reported low to very low self-perceived knowledge about COVID-19 vaccines. Only 5.8% of the students have been vaccinated for influenza in 2019 and 2020. The 43.8% of participants somewhat or completely agreed to accept a safe and effective COVID-19 vaccine, while 22.2% somewhat or completely disagreed to accept this vaccine. Trust in government, doctors regarding the information about the COVID-19, and government experts regarding the information about the COVID-19 was 35.5%, 71.4% and 63.8% respectively. The most important reason for refusal of a COVID-19 vaccine was doubts about the safety, efficacy and effectiveness of the vaccine (72.4%). Mean score on fear of COVID-19 scale was 14.7 (standard deviation=5.9), while the median score was 14 (range=28).

**Table 2.** Students’ answers regarding the COVID-19 and vaccination.

|  | N | % |
| --- | --- | --- |
| Contact with a confirmed or a suspected case of COVID-19 |  |  |
| Yes | 1142 | 50.9 |
| No | 1103 | 49.1 |
| Infected with COVID-19 |  |  |
| Yes | 266 | 11.8 |
| No | 1981 | 88.2 |
| Family/friends infected with COVID-19 |  |  |
| Yes | 1354 | 60.4 |
| No | 888 | 39.6 |
| Self-perceived likelihood of getting infected with the COVID-19 in the future |  |  |
| Very low | 80 | 3.8 |
| Low | 470 | 22.4 |
| Moderate | 999 | 47.7 |
| High | 397 | 18.9 |
| Very high | 150 | 7.2 |
| Self-perceived knowledge about the COVID-19 |  |  |
| Very low | 16 | 0.7 |
| Low | 63 | 2.8 |
| Moderate | 820 | 36.5 |
| High | 1070 | 47.6 |
| Very high | 279 | 12.4 |
| Self-perceived knowledge about COVID-19 vaccines |  |  |
| Very low | 403 | 17.9 |
| Low | 833 | 37.0 |
| Moderate | 800 | 35.6 |
| High | 161 | 7.2 |
| Very high | 52 | 2.3 |
| Influenza vaccination in 2019 and 2020 |  |  |
| Yes | 130 | 5.8 |
| No | 2114 | 94.2 |
| Accept a safe and effective COVID-19 vaccine |  |  |
| Completely disagree | 269 | 12.0 |
| Somewhat disagree | 230 | 10.2 |
| Neutral | 765 | 34.0 |
| Somewhat agree | 545 | 24.2 |
| Completely agree | 440 | 19.6 |
| Trust in government |  |  |
| Yes | 794 | 35.5 |
| No | 1445 | 64.5 |
| Trust in doctors regarding the information about the COVID-19 |  |  |
| Yes | 1604 | 71.4 |
| No | 641 | 28.6 |
| Trust in government experts regarding the information about the COVID-19 |  |  |
| Yes | 1432 | 63.8 |
| No | 814 | 36.2 |
| Reasons for refusal of a COVID-19 vaccine |  |  |
| I have doubts about the safety, efficacy and effectiveness of the vaccine | 1129 | 72.4 |
| I believe that even if I get infected with COVID-19, nothing bad will happen to me | 99 | 6.3 |
| I believe that the vaccine is not necessary | 84 | 5.4 |
| I do not believe in the necessity of vaccines | 76 | 4.9 |
| I believe that I will not be infected by COVID-19 | 35 | 2.2 |
| I believe that the COVID-19 virus is not particularly dangerous | 29 | 1.3 |

Students from Italy gave the highest proportion of positive responses (“somewhat agree” and “completely agree”) regarding COVID-19 vaccination (121 of 170 students, 71.2%), and then students from Spain (117 of 181, 64.6%), Greece (161 of 275, 58.5%), Cyprus (57 of 131, 43.5%), Kosovo (393 of 1020, 38.5%), Albania (102 of 313, 32.6%) and Czech Republic (34 of 159, 21.4%).

Univariate and multivariate logistic regression analysis with intention to accept a safe and effective COVID-19 vaccine as the dependent variable is shown in Table 3. Multivariable analysis showed that males (OR=1.41, p=0.008) and students that did not work in healthcare facilities during the COVID-19 pandemic (OR=1.58, p=0.001) were more willing to accept COVID-19 vaccination. In addition, students that took influenza vaccination in 2019 and 2020 were more amenable to take COVID-19 vaccine also (OR=2.38, p<0.001). Trust in government (OR=1.85, p<0.001), in doctors regarding the information about the COVID-19 (OR=2.13, p<0.001), and in government experts regarding the information about the COVID-19 (OR=1.40, p=0.012) increased the probability of getting vaccinated with regards to COVID-19. Increased self-perceived knowledge about COVID-19 vaccines (moderate vs. very low/low; OR=1.39, p=0.001, high/very high vs. very low/low; OR=1.86, p<0.001), and increased fear of COVID-19 were related with increased likelihood of COVID-19 vaccination (low vs. very low; OR=1.73, p<0.001, medium vs. very low; OR=2.02, p<0.001, high vs. very low; OR=2.15, p<0.001, very high vs. very low; OR=1.75, p<0.001).

**Table 3.**
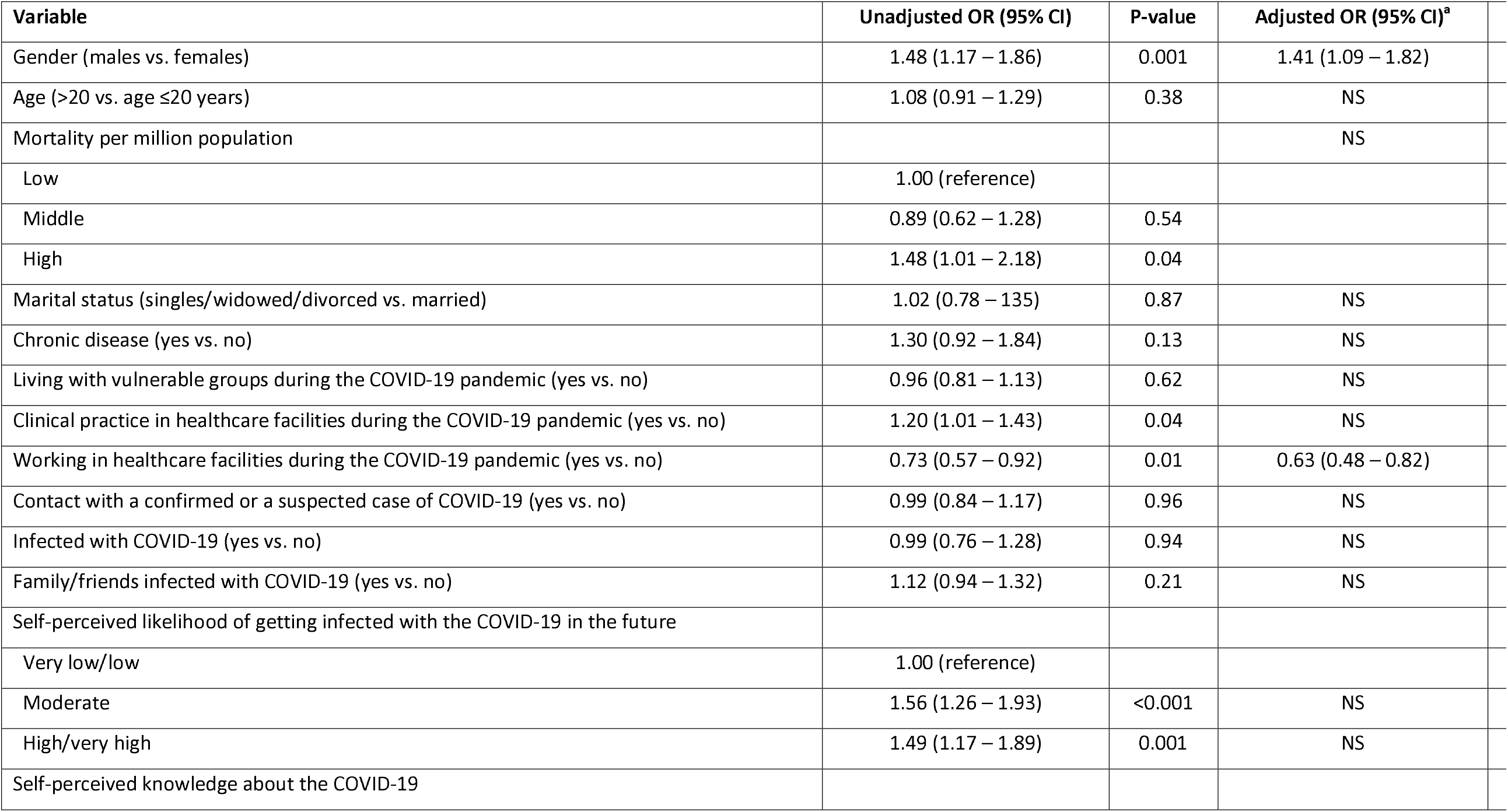
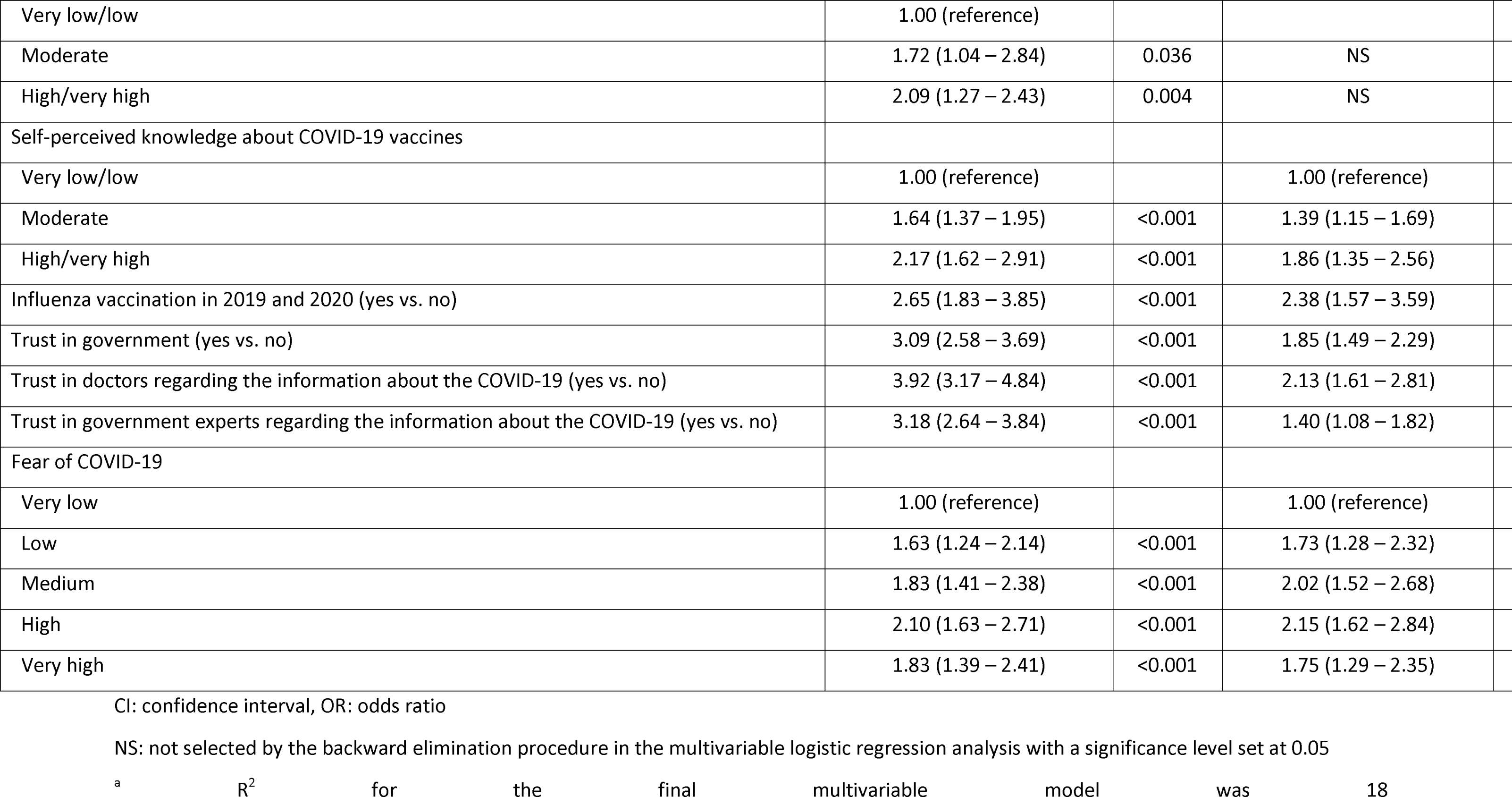
Univariate and multivariate logistic regression analysis with intention to accept a safe and effective COVID-19 vaccine as the dependent variable.

| Variable | Unadjusted OR (95% CI) | P-value | Adjusted OR (95% CI)<sup>a</sup> |
| --- | --- | --- | --- |
| Gender (males vs. females) | 1.48 (1.17 – 1.86) | 0.001 | 1.41 (1.09 – 1.82) |
| Age (>20 vs. age ≤20 years) | 1.08 (0.91 – 1.29) | 0.38 | NS |
| Mortality per million population |  |  | NS |
| Low | 1.00 (reference) |  |  |
| Middle | 0.89 (0.62 – 1.28) | 0.54 |  |
| High | 1.48 (1.01 – 2.18) | 0.04 |  |
| Marital status (singles/widowed/divorced vs. married) | 1.02 (0.78 – 1.35) | 0.87 | NS |
| Chronic disease (yes vs. no) | 1.30 (0.92 – 1.84) | 0.13 | NS |
| Living with vulnerable groups during the COVID-19 pandemic (yes vs. no) | 0.96 (0.81 – 1.13) | 0.62 | NS |
| Clinical practice in healthcare facilities during the COVID-19 pandemic (yes vs. no) | 1.20 (1.01 – 1.43) | 0.04 | NS |
| Working in healthcare facilities during the COVID-19 pandemic (yes vs. no) | 0.73 (0.57 – 0.92) | 0.01 | 0.63 (0.48 – 0.82) |
| Contact with a confirmed or a suspected case of COVID-19 (yes vs. no) | 0.99 (0.84 – 1.17) | 0.96 | NS |
| Infected with COVID-19 (yes vs. no) | 0.99 (0.76 – 1.28) | 0.94 | NS |
| Family/friends infected with COVID-19 (yes vs. no) | 1.12 (0.94 – 1.32) | 0.21 | NS |
| Self-perceived likelihood of getting infected with the COVID-19 in the future |  |  |  |
| Very low/low | 1.00 (reference) |  |  |
| Moderate | 1.56 (1.26 – 1.93) | <0.001 | NS |
| High/very high | 1.49 (1.17 – 1.89) | 0.001 | NS |
| Self-perceived knowledge about the COVID-19 |  |  |  |
| Very low/low | 1.00 (reference) |  |  |
| Moderate | 1.72 (1.04 – 2.84) | 0.036 | NS |
| High/very high | 2.09 (1.27 – 2.43) | 0.004 | NS |
| Self-perceived knowledge about COVID-19 vaccines |  |  |  |
| Very low/low | 1.00 (reference) |  | 1.00 (reference) |
| Moderate | 1.64 (1.37 – 1.95) | <0.001 | 1.39 (1.15 – 1.69) |
| High/very high | 2.17 (1.62 – 2.91) | <0.001 | 1.86 (1.35 – 2.56) |
| Influenza vaccination in 2019 and 2020 (yes vs. no) | 2.65 (1.83 – 3.85) | <0.001 | 2.38 (1.57 – 3.59) |
| Trust in government (yes vs. no) | 3.09 (2.58 – 3.69) | <0.001 | 1.85 (1.49 – 2.29) |
| Trust in doctors regarding the information about the COVID-19 (yes vs. no) | 3.92 (3.17 – 4.84) | <0.001 | 2.13 (1.61 – 2.81) |
| Trust in government experts regarding the information about the COVID-19 (yes vs. no) | 3.18 (2.64 – 3.84) | <0.001 | 1.40 (1.08 – 1.82) |
| Fear of COVID-19 |  |  |  |
| Very low | 1.00 (reference) |  | 1.00 (reference) |
| Low | 1.63 (1.24 – 2.14) | <0.001 | 1.73 (1.28 – 2.32) |
| Medium | 1.83 (1.41 – 2.38) | <0.001 | 2.02 (1.52 – 2.68) |
| High | 2.10 (1.63 – 2.71) | <0.001 | 2.15 (1.62 – 2.84) |
| Very high | 1.83 (1.39 – 2.41) | <0.001 | 1.75 (1.29 – 2.35) |

CI: confidence interval, OR: odds ratio
NS: not selected by the backward elimination procedure in the multivariable logistic regression analysis with a significance level set at 0.05
<sup>a</sup> R<sup>2</sup> for the final multivariable model was 18

## Discussion

According to our best knowledge, this is the first study worldwide that attempts to explore the intentions of nursing students to get vaccinated for SARS-CoV-2 infection in seven European countries. With several vaccines having been approved by the respective agencies in USA, Europe and other countries the main challenge now for health policy makers worldwide is the acceptance and vaccination of the population. According to the results of the current study, key reasons for willingness to get vaccinated were male gender, having not worked in healthcare facilities during the pandemic, getting vaccinated for influenza in 2019 and 2020, trust in doctors, governments and experts and higher level of knowledge and fear about COVID-19.

Less than half of the study participants reported that would get vaccinated if a vaccine results as safe and effective. We expected acceptance rates of a safe and effective vaccine could be higher among nursing students. Due to their future profession, they have more knowledge about benefits of vaccines and are more awareness about their need. In a study that evaluated the attitude of population regarding future vaccination, half of them (49.7%) reported a positive willingness with students and healthcare personnel being more willingness in comparison to other occupational groups [17]. An Italian study among students concluded that 86.1% are willing to get vaccinated for COVID-19 [18] while a study in Malta among healthcare students reported acceptance rates at a level of 44.2% [19]. A study among medical students reported willingness to receive a vaccine after getting approval at 77% [20]. Differences in the sample, data collection, morbidity and mortality rates due to COVID-19 and different periods of studies conduction are most probably the reasons for these inconsistences. Receiving information from non-trusted sources has most probably an impact in the low rates. In a study among college students in South Carolina, only 57.7% received information about vaccines from health agencies with social media and personal networks being also the two other main sources [21]. Another study among medical and healthcare students report that social media are a key source for them to receive information about COVID-19 [22].

However, in the current study most of the participants who refuse to get vaccinated reported as the main reason the doubts about safety, efficacy and effectiveness. Other reasons for not getting vaccinated were reported the beliefs that even if they get infected nothing bad will happen, beliefs that vaccines are not necessary, beliefs that will not get infected with the virus etc.

Men are more likely to get vaccinated than women. Similar results are presented also in other studies with women being more reluctant to get vaccinated in comparison to men [17, 23]. Biological and lifestyle factors and immune response differs among men and women. In general, women take into consideration COVID-19 more seriously and respect more the prevention and public health rules [24]. In contrast, female students are more likely to believe in conspiracy theories [25]. However, even though prevalence of the disease is similar to both sexes, the outcome and death rates are higher for men than women [26]. Most probably, this makes women more confident for the outcome and more reluctant to get vaccinated, but this should be interpreted with caution.

Participants who have worked in healthcare facilities are less positive in getting vaccinated. The intention of getting vaccinated for COVID-19 ranged from 50% - 93.3% (depending on vaccine effectiveness) among Indonesian healthcare personnel [27]. A study among Maltese healthcare personnel reaches the conclusion that around half of them (52%) are intending to get vaccinated [28] while another study in USA reported levels of 57.6% [29]. A meta-analysis showed that healthcare personnel intention for getting vaccinated for COVID-19 ranges from 43.6%-67.9% [30]. People who work in healthcare facilities usually are in higher risk of getting infected, but this doesn’t seem to make them more aware about this need. A study among healthcare workers in China shows that the high possibility of getting infected increase the chances of vaccination [31]. Vaccination rates for influenza have been reported low in nurses population [32, 33]. Additionally, a study has reported a negative connection between nursing profession and vaccination [34]. Focusing on this population to get vaccinated is of paramount significance and specific strategies should be developed from healthcare authorities at both national and local level. Having a healthy future healthcare personnel can contribute in the better management of the pandemic but also can serve as a role model for general population to follow the same attitude.

The attitude toward general vaccination contributes significantly in vaccine hesitancy [35]. In our study, students that have been vaccinated for common influenza in 2019 and 2020 were more likely to have intentions to get vaccinated with a safe and effective COVID-19 vaccine. Similar results have been reported in different studies among students [19] and healthcare personnel [28]. Despite the well-established knowledge about benefits of influenza vaccination, the rates among nursing students remain low [36-38]. Development of vaccine policies, organization of campaigns for raising awareness of students and provision of vaccine out of charge could help in increasing the vaccination rates.

According to the results of the current study, people who trust the governments, the experts and the doctors were more willingness to get vaccinated. In a recent study conducted in 19 countries, people that trusted information provided from government sources have higher probability to accept a COVID-19 vaccine [39]. Students who trust the public health experts were more likely to get vaccinates was reported in another study [20]. Receiving information from scientists was significantly correlated with high vaccine acceptance [21]. The last period different conspiracy theories about the virus, the public health measures and the vaccines exist. People who tend to believe these theories are more likely to be resistant to preventive measures and vaccination [40]. A study concluded that people who believe in conspiracy theories have 3.9 times less intention not to get vaccinated and not to support the public health preventive measures [41]. In general, people tend to believe more experts and doctors than politicians. It is of paramount significant to identify those who tend to believe these theories and to provide reliable information. Political polarization and disputes between political parties have decreased also public trust [42]. Doctors can play an important role in enhancing trust and addressing conspiracy theories [41]. Vaccine recommendation from a healthcare provider increase the chances of getting vaccinated [43]. Collaboration and synergies between policymakers, stakeholders, civil society and local communities could help in trust establishment. Additionally, a minimum consensus from political parties in a bipartisan management of the pandemic could also help.

Participants with higher level of knowledge about COVID-19 vaccines were more likely to get vaccinated in contrast to those with low level. Studies have shown that students with high level of vaccine knowledge are more likely to vaccinate themselves [44, 45]. Increasing the knowledge of target population about vaccines could improve uptake rates. Additionally, inclusion of the vaccination benefits should be a regular part for healthcare professions curricula [46].

Participants who were more feared about COVID-19 were more likely to get vaccinated. Similar results are reported in a study among college students in USA [35] and general population [23]. Level of fear have been reported high in different studies among University students [47]. In general, mental health status of nursing students has deteriorated during the pandemic. Many studies have reported high level of depression and anxiety during the COVID-19 period [4, 48]. Public health emergencies have a significant impact on student’s mental health status. Provision of support by authorities (governmental and universities) could decrease these symptoms [49].

## Strengths and limitations

Our study suffers from some limitations. This was a cross-sectional study and extraction of causalities is difficult. Additionally, the sample is not totally representative of the seven countries that participated mainly due to the on-line data collection method. In this study participated only nursing students and generalization of the results to other students is difficult. Also, self-reported questionnaires could always result on information bias.

However, to our best knowledge, this is the first study that assessed factors that influence nursing students’ intention to accept COVID-19 vaccination in seven European countries with a large geographic coverage. This study identifies key factors that should be addressed in order to increase COVID-19 vaccine uptake rates. These results could be very useful and helpful for policy makers who develop vaccine promotion strategies.

## Conclusions

In the current study, 2249 nursing students participated from seven European countries. The results of the current research concluded that key factors for intending to get vaccinated were male gender, not having worked in healthcare facilities during the pandemic, being vaccinated for influenza in 2019 and 2020, trusting doctors, governments and experts, having high level of knowledge and fear about COVID-19. Doubts about safety, efficacy and effectiveness was reported by the participants as the main reason of not willing to vaccinate.

Implementation of vaccination policies, organization of awareness campaigns and increasing knowledge about vaccine benefits are some key recommendations to be followed. Synergies and collaboration between policymakers and university authorities is of paramount significance. Politicians should be less involved in vaccination campaigns with doctors and healthcare experts having the main role. Regular assessment of mental health status and provision of support could decrease the fear levels for this population.

## Data Availability

Data will be available after request.

## Funding

This research did not receive any specific grant from funding agencies in the public, commercial, or not-for-profit sectors.

## Declaration of Competing Interest

There is no conflict of interest.

## Acknowledgements

The authors warmly thank all the nursing students who participated in the current study.

## Notes

### Competing Interest Statement

The authors have declared no competing interest.

### Funding Statement

None to declare

### Author Declarations

Ethical approval from the Research and Ethics Committee of the Hellenic Mediterranean University 63-29.11.21.

